# Association Between Cannabis Smoking and Academic Achievement in Colombian High-School Students

**DOI:** 10.1101/2022.02.22.22271351

**Authors:** Yegson Pérez-Martínez, Guillermo Augusto Ceballos-Ospino, Adalberto Campo-Arias

**Author notes:** **Author note** Correspondence to Adalberto Campo-Arias, Programa de Medicina, Facultad Ciencias de la Salud, Universidad del Magdalena, Carrera 32 No 22-08, Santa Marta, Colombia (código postal 470004). Teléfono: 57 5 438100, extensión 1338. The Research Vice-rectory of the University of Magdalena financed this research through Resolution 0347 of 2018 in the name of Adalberto Campo-Arias (Call Fonciencias 2017). The authors have no conflicts of interest to declare. **Contributors:** Yegson Pérez-Martínez contributed to the study conception and data interpretation, and statistical analysis, drafted the article, and revised and approved the final version. G. A. Ceballos-Ospino and A. Campo-Arias contributed to the study design and conception, data interpretation, statistical analysis, revised the intellectual content and approved the final version.

## Abstract

**Background:** Cannabis smoking can affect academic achievement, and depressive symptoms and family dysfunction are also associated it. The study’s objective was to establish the relationship between cannabis smoking and academic achievement, controlling clinically important depressive symptoms and family dysfunction.

**Methods:** A cross-sectional study was designed. The authors quantified lifetime cannabis smoking, perception of academic achievement, clinically important depressive symptoms, and family dysfunction. The crude association between cannabis smoking and academic achievement was computed; after it was adjusted by clinically important depressive symptoms and family dysfunction.

**Results:** 1,462 students between 13 and 17 participated in the research; 11.6% reported lifetime cannabis smoking, 30.8% poor-fair academic achievement; 7.1% clinically important depressive symptoms; and 76.1% family dysfunction. Lifetime cannabis smoking was significantly associated with poor-fair academic achievement after adjusting for clinically important depressive symptoms and family dysfunction (OR = 1.61, 95%CI 1.16 - 2.24).

**Conclusions:** Lifetime cannabis smoking is related to poor-fair academic achievement among high-school students in Santa Marta, Colombia.

**Data availability statement:** The data supporting this study’s findings are available from the corresponding author upon reasonable request.

**Authors biography:** Yegson Pérez-Martínez

He is a young psychologist interested in researching school psychology, academic achievement, and health-compromising behaviours.

Guillermo Augusto Ceballos-Ospino, psychologist He is a psychologist and occasional professor at the Program of Psychology, University of Magdalena, Santa Marta, Colombia. His action areas are mental health, especially suicide prevention, and psychometrics. He is interested in studies with populations of university students.

Adalberto Campo-Arias, MD, MSc He is a psychiatrist, magister in sexual and reproductive health and professor at the School of Medicine, Faculty of Health Science, University of Magdalena, Santa Marta, Colombia. His research interests include sexual and reproductive health, human rights, prejudice and discrimination, and health-compromising behaviours.

## 1. INTRODUCTION

In 2016, cannabis smoking increased worldwide, and it was the most consumed drug; around 192 million people used it at least once. Cannabis is the substance most used by young people due to easy access and is mistakenly considered a low health risk. Cannabis smoking usually begins in adolescence and is often the gateway to the use of other illegal substances [1].

In Latin America and the Caribbean, the frequency of cannabis smoking in secondary school students follows the world trend in countries such as Argentina, Barbuda, Chile, and the Dominican Republic, with prevalence close to 20%. However, there are essential discrepancies in the cannabis use frequency because in countries such as Colombia and Ecuador, a prevalence of less than 10% was documented [2]

### 1.1. Cannabis smoking

Cannabis smoking in adolescents is related to different risk factors: individual, peer, family, community, and society [3]. At the individual level, cannabis smoking is associated with adjustments typical of the normative period of adolescence, the need to consolidate or reaffirm an identity, curiosity about new experiences, and misinformation [4]. It is hypothesized that adolescents use cannabis to seek approval and demonstrate competence [5].

Among the family factors, the poor communication between father and children, poor parenting styles, parental absenteeism, and a history of substance use in the family group stand out. In the group of community factors, the availability of time, access to the substance in the sector, and the advertising that reaffirms the need to try new things can be pointed out [6].

### 1.2. Academic achievement

Academic achievement cannot be reduced to simply understanding students’ attitudes and aptitudes [7]. Academic achievement also implies fulfilling the courses’ goals, achievements, and objectives and the knowledge shown in the subjects studied in qualitative and quantitative measurements [8].

Different factors influence adolescent academic achievement, some personal and others associated with the family and school context [7]. Personal factors characterize the student as a learner [9]. The multiple academic objectives and goals that intrinsically motivate the student in academic settings should be considered [8].

The contextual factors refer to the family, social, economic status, and the school environment where the student develops [8]. Likewise, contextual factors may include the relevance of the perception of performance by parents. This aspect is crucial because it can become a stressor that mediates the student’s psychological well-being and self-concept. Consequently, it can be affirmed that parents can directly or indirectly influence academic achievement [7].

### 1.3. Association between cannabis smoking and academic achievement

Cannabis smoking can unfavourably affect academic achievement; Hayatbakhsh and colleagues [10], in a sample of 3,478 Australian students, found that frequent cannabis smoking increased almost twice the risk of low academic achievement. Cox and colleagues [11] found in 1,488 high school students from the United States that cannabis smoking was two-fold associated with low academic achievement. Finally, Guerrero-Martelo and colleagues [12], in 156 Colombian students, found that cannabis smoking increased the risk of low academic achievement ten times.

#### 1.3.1. Confounding variables in the relationship between cannabis smoking and academic achievement: depressive symptoms and family dysfunction

##### 1.3.1.1. Depressive symptoms, cannabis smoking, and academic achievement

It should be considered that the association between cannabis smoking and academic achievement is usually negatively modified by other variables such as depressive symptoms and family functioning [13-16].

First, it should be considered that cannabis smoking can be associated with depressive symptoms and be a confusing variable in the association between cannabis smoking and academic achievement since depressive symptoms can independently affect academic achievement unfavourably. Rojas and colleagues [17] found that in a sample of 2,597 Chilean students that cannabis smoking increased two times the risk of depression.

In the same way, it is observed that depressive symptoms can negatively affect academic achievement. In Colombia, Campo-Arias and colleagues [13], in a sample of 560 students from Bucaramanga, observed that depressive symptoms with clinical importance were related at least two-fold to low academic achievement. However, Cogollo and colleagues [14], in a sample of 512 students from a public school in Cartagena, documented that depressive symptoms did not maintain a significant relationship with academic achievement.

##### 1.3.1.2 Family functioning, cannabis smoking, and academic achievement

Another variable that can qualify the relationship between cannabis smoking and academic achievement is family functioning; there is empirical evidence that supports the association in different sociocultural contexts. Zarrouq and colleagues [16], in 3,020 students from Morocco between 11 and 23 years old, found that feeling insecure with the family, one of the components measured for family dysfunction was significantly associated two-fold with substance use psychoactive.

Likewise, a statistically significant association between family dysfunction and low academic achievement has been observed. Gómez-Bustamante and colleagues [15], in 1,730 students from Cartagena, Colombia, between 13 and 17 years, found that family dysfunction was almost two times related to low academic achievement.

The studies’ limitations are common in the association between cannabis smoking, academic achievement, depressive symptoms, and family dysfunction. These variables can act as confounding variables or interact with academic achievement [18]. The studies that analyzed the association between cannabis smoking and academic achievement did not adjust for the relationship with confounding variables, such as depressive symptoms and family function [11, 12]. The present research studied and adjusted the association between cannabis smoking and academic achievement for depressive symptoms and family function.

### 1.4. Theoretical bases of the association

The use of substances, such as cannabis, is a learned behaviour that can be modified and extinguished. A behaviour from which pleasure and approval are obtained tends to repeat itself; it can be configured as habitual behaviour. If the behaviour does not get satisfying results, it is less likely to repeat, and consequently, the probability of becoming a habit is reduced [7].

The association between cannabis smoking and academic achievement can be explained by the neurological damage induced by cannabis [19]. This theory explains that cannabis smoking directly affects the central nervous system’s functioning, mainly on synaptic function related to neurotransmitters [20]. Delta-9-tetrahydrocannabinol and endogenous cannabinoids, such as anandamide, are related to repeated consumption patterns that can inhibit the production or block the reception of neurotransmitters in the synaptic cleft [19]. These changes directly affect higher mental functions, such as information processing, reflective thinking, memory, motor activity, and visual perception [19, 20]. The integrity of functions is necessary for the school setting to achieve excellent academic achievement [19].

The theory of cognitive distortions can convincingly explain the relationship between depressive symptoms and academic achievement [21]. Negative emotional states such as depression generate errors in information processing; such distortions can manifest as disruptive behaviours or low academic achievement [22]. In this way, people with depressive symptoms present what is known as the negative cognitive triad, which causes the individual to have a negative view of himself, the world, and the future, which can be reflected in little interest in achieving goals such as excellent academic achievement. The cognitive distortions associated with depressive symptoms affect labelling, thinking, overgeneralization, guilt, catastrophizing, and emotional reasoning [21]. These alterations limit the learning process, reduce motivation, and interfere with intellectual activity, processes directly involved in academic achievement [23].

Bertalanffy’s general system theory explains the relationship between family function and academic achievement [24]. This theory explains the organization of various phenomena in which different variables or actors participate. Human development is framed in different interaction scenarios with the environment ecological theory. This postulate maintains that the family is the system that broadly defines and configures the development of the person, in which it is affirmed as systems, microsystems, areas close to the individual (family, school, peer group, the neighbourhood), mesosystems (interaction of two or more microsystems), macrosystems (culture, and ideologies), directly influence or affect development in the individual [25].

Human development is framed and congregates from the influence of different family systems, schools, peers, and cultures, all related to each other. Likewise, if any of these systems are dysfunctional, they can negatively impact each individual’s development, which makes it up [25]. Substance use, family dysfunction, and depressive symptoms, only in conjunction or interaction, affect academic performance and can be indicators of problems in the systems in which students operate due to their dynamic structure [24].

Depressive symptoms are associated with cognitive distortions [21, 22]; they can minimize the risk of cannabis smoking [25]. Likewise, it should be remembered that depressive episodes manifest as difficulty concentrating on academic work and decreased self-efficacy [26, 27]. Also, it is necessary to remember that cannabis smoking can be a form of self-medication to face or escape the stress associated with family function or the discomfort typical of depressive symptoms. Stress and cannabis smoking can have a negative brain effect in the short and long term on higher mental functions that can explain low academic achievement [19, 20].

### 1.5. Practical implications

Thanks to the knowledge of the variables that influence behaviour acquisition and the fundamental role of reinforcers. Strategies can be established to modify behaviours that generate a risk to the health of individuals, such as consumption; if the reduction of reinforcers that maintain these behaviours is changed and if they enhance those protective factors, more acceptable social behaviours can be achieved [18].

The present cross-sectional study’s findings may contribute to the recognition that low academic achievement is not only due to factors intrinsic to students, but different contextual elements can influence, directly or indirectly, and condition academic achievement [9]. This knowledge allows us to justify the creation of programs for teachers. Educators have the role of guiding and changing agents to decrease cannabis smoking [6, 28]. Furthermore, the findings suggest working in an interdisciplinary way to address the risk factors and identify possible cases of significant cannabis smoking or depressive symptoms. These strategies can favour academic achievement and reduce depressive symptoms within the family system [6, 28, 29].

The family plays a fundamental role from childhood in the foundation of the personality bases. Based on the fact that the family is the social institution that integrates the factors, it is previously noted that the integral formation of children and adolescents [29]. Most habits and customs maintained throughout life are initiated and consolidated [30]. Proper family functioning may be necessary to reduce the initiation and maintenance of cannabis smoking and, at the same time, to achieve the best achievements in school [31].

This study’s objective was to estimate the association between cannabis smoking and academic achievement, adjusted by depressive symptoms and family function, in high school students from Santa Marta, Colombia.

## 2. METHOD

### 2.1. Design and ethical considerations

It was designed as a cross-sectional study. Ethical standards for research involving humans were followed according to Colombian laws and the Helsinki declaration: The project was reviewed and approved by the Research Ethical Board of the Universidad del Magdalena (an ordinary session on July 12, 2018), parents or guardians signed informed consent, and student’s consented to participate in the research.

### 2.2. Population and sample

A probabilistic sampling was followed in several stages based on clusters. The size of each cluster was an estimated 25 students. This sample size allows working with prevalence between 3% (± 1) and 50 (± 5), with a confidence level of 95%. Additionally, a 20% replacement was calculated to cover possible losses due to different eventualities, such as denying educational institutions’ authority, not having the parents’ informed consent, or denying student assent. Thus, a sample of 1,948 students was projected. This sample size would also allow associations to be explored and adjusted with relatively narrow 95% confidence intervals. Students between the ages of 13 and 17 with the skills to fill out the research booklet were included.

### 2.3. Instruments

#### 2.3.1. Cannabis smoking

The cannabis smoking in the life course was quantified with one item taken from the Youth Risk Behavior Survey Questionnaire, epidemiological surveillance of risk behaviours in middle-school students in the United States: Have you ever used marijuana? The item presents a dichotomous response option [32]. This questionnaire has acceptable reproducibility in-school adolescents [33].

#### 2.3.2. Academic achievement

During the last month, the perception of academic achievement was evaluated with an item. This item presents four response options: excellent, good, fair, and poor. For the present analysis, the responses were dichotomized into excellent/good and poor/fair.

#### 2.3.3. Depressive symptoms

Depressive symptoms were quantified with the World Health Organization General Well-being Index (WHO-5). The WHO-5 has five items that address the perception of general well-being: the absence of depressive symptoms during the last fifteen days: good mood, tranquillity, feeling energetic, quality of sleep, and interest in daily life activities. Each item has four response options: never, sometimes, many times, and always. Each response is scored from zero to three. Consequently, the total scores are between zero and fifteen; the lower the score, the more depressive symptoms [34].

In the present study, scores were dichotomized into depressive symptoms without and with clinical significance, and scores of 8 or less were categorized as depressive symptoms with clinical significance.

#### 2.3.4. Family functioning

The family function was quantified with the family APGAR. This questionnaire has five items that address the family group’s adaptability, cooperation, development, affectivity, and resolution capacity. Each item gives five response options: Never, seldom, sometimes, almost always, and always. The options are scored between zero and four; therefore, the total scores range between zero and twenty [35]. In the current study, the scores were dichotomized, and those fifteen or fewer were categorized as family dysfunction.

### 2.4. Procedure

The information collection process was carried out between September 1 and October 31, 2018. The students completed the questionnaire as a group on an ordinary day in the classroom. In the classroom, the research assistants’ team explained the research’s objectives, the importance of anonymizing the name to fill out the item booklet, and the general response rules. In the same way, the group of research assistants offered to clarify doubts without inducing responses.

### 2.5. Analysis of data

Cannabis smoking was the independent variable, academic achievement was the dependent variable, and family function and depressive symptoms as confounding variables. First, crude odds ratios (OR) with a 95% confidence interval (95% CI) were estimated; after, adjusted OR for the family function and depressive symptoms was computed. The analysis was performed using the IBM-SPSS program, version 22.

## 3. RESULTS

A total of 1,462 students participated in the research, the ages were between 13 and 17 (M = 16.0, SD = 0.8). The sociodemographic description of the student sample is presented in Table 1.

**Table 1.** Demographic characteristics of the high-school students.

| Variable | n | % |
| --- | --- | --- |
| Age (years) |  |  |
| 13 - 15 | 425 | 29.1 |
| 16 - 17 | 1,037 | 70.9 |
| **Gender** |  |  |
| Female | 882 | 60.3 |
| Male | 580 | 39.7 |
| **Grade** |  |  |
| Tenth | 809 | 55.3 |
| Eleventh | 653 | 44.7 |
| **Income** |  |  |
| Low | 725 | 49.6 |
| Middle | 508 | 34.7 |
| High | 39 | 2.6 |
| No answer | 190 | 13.0 |

A total of 170 students (11.6%) reported cannabis smoking; 450 students (30.8%), poor-fair academic achievement; 103 students (7.1%), depressive symptoms; and 1,112 students (76.1%), family dysfunction. The WHO-5 presented a Cronbach’s alpha of 0.82, and the family APGAR showed a Cronbach’s alpha of 0.81.

Cannabis smoking was significantly associated with poor-fair academic achievement (OR = 1.73, 95%CI 1.25 - 2.40). Even after adjusting for depressive symptoms and family dysfunction (OR = 1.61, 95%CI 1.16 - 2.24).

## 4. DISCUSSION

The present study shows the association between cannabis smoking and academic achievement; after controlling for depressive symptoms and family dysfunction, a significant association was maintained with the variables adjusted for poor-fair academic achievement. This finding, OR = 1.61, 95%CI 1.16 - 2.24, is similar to other cross-sectional studies that found a statistically significant association between cannabis smoking and low academic achievement. In Australia, Hayatbakhsh and colleagues [10] reported that cannabis smoking raises the odds of low academic achievement (OR = 1.70, 95%CI 1.10 - 2.80). In the United States of America, Cox and colleagues [11] documented that cannabis smoking was related to low academic achievement (OR = 1.90, 95%CI 1.20 - 2.90). Similarly, in a small sample of Colombian students, Guerrero-Martelo and colleagues [12] found that cannabis smoking raises the possibility of low academic achievement (OR = 11.00, 95%CI 3.53 - 37.43). Some longitudinal studies showed that cannabis smoking increased academic dropout risk; academic dropout is generally associated with poor academic achievement [36]. Likewise, it was observed that the age of onset of cannabis smoking was associated with low academic achievement and reduced the chance of graduating [37]. Adolescents who started cannabis smoking earlier showed lower academic achievement than those who started cannabis smoking at an older age [38]. However, in a study with participants with a history of cannabis smoking, it was observed that cannabis smoking was not associated with a missing school or repeating the last year [39]. Similarly, no statistically significant association was found between cannabis smoking and academic achievement after controlling cigarette smoking and alcohol consumption [40].

Given the study’s cross-sectional design, it might be thought that low academic achievement could increase cannabis smoking. In agreement with this hypothesis, Hayatbakhsh and colleagues [10] observed, in a longitudinal study, that habitual cannabis smoking increased the risk of low academic achievement. However, in a cohort study, Fergusson and colleagues [36] did not observe that low academic achievement increased the risk of cannabis smoking.

Early and prolonged exposure to cannabis smoking can cause significant alterations in higher cognitive functions due to functional and structural alterations at the brain level [20]. Depending on the use pattern, cannabis negatively affects attention, memory, and learning up to weeks after the acute effects of the substance [19]. However, in the present study, lifetime cannabis smoking was only quantified without investigating the age of onset and frequency of use. It is possible that in adolescents, cannabis smoking can be a form of self-medication to face negative emotional states, such as depressive symptoms and those associated with family dysfunction, and thus affect academic achievement [15]. Family dysfunction and depressive symptoms can also negatively affect academic achievement [13-16]. Hence, adjusting simultaneously for these confounding variables is necessary [18].

Depressive symptoms are associated with errors in information processing and cognitive distortions such as labelling, polarized thinking, overgeneralization, guilt, catastrophization, and emotional reasoning [21, 22]. These changes reduce self-efficacy motivation and hinder learning, reducing academic achievement [23].

Family dysfunction is highly prevalent in Colombian families with adolescent children [14, 15]. Likewise, family dysfunction can be considered an essential stressor in the student’s life that disturbs emotional well-being and can deteriorate academic achievement [15]. It is necessary to control family function in Colombian studies investigating variables associated with academic performance.

### 4.1. Significance for public health

Academic achievement is the result of intrinsic and extrinsic variables [8]. The design of the present study suggests that cannabis smoking can impair academic achievement, but, in turn, depressive symptoms and family dysfunction can increase the risk of both cannabis smoking and deteriorate academic achievement [10, 11, 13-16]. Consequently, low academic achievement cases must be studied comprehensively and consider many variables [14, 15].

### 4.2. Strengths and limitations

Thus, this research’s critical contribution was to analyze the relationship between cannabis smoking and academic achievement after adjusting depressive symptoms and family function, previously not considered confounding variables in the association. However, this study has limitations; it is impossible to establish causal relationships between the variables because these are simultaneously measured [18]. Besides, cannabis smoking frequency was not established. The severity of consumption is directly related to a negative impact at the cognitive level and, consequently, on academic performance [19, 20]. Finally, academic performance was investigated by self-report, and there was no objective and quantitative data on academic performance.

## 4.3. Conclusions

It is concluded that cannabis smoking is significantly associated with poor-fair academic achievement after adjusting for depressive symptoms and family dysfunction in high school students from Santa Marta, Colombia. Low academic achievement must be studied comprehensively and consider many variables, including substance use, such as cannabis. Personal variables such as the presence of depressive symptoms and contextual variables such as family functioning should always be considered in the relationship between cannabis use and academic performance. Moreover, researchers should consider the possible bidirectional relationship between substance use and academic performance in adolescent students. Further research and consideration of other confounding variables are needed.

## Data Availability

The data supporting this study's findings are available from the corresponding author upon reasonable request.

